# Estimating the Burden of COVID-19 Symptoms Among Participants at the 2020 USA Curling Club Nationals Tournament

**DOI:** 10.1101/2020.10.08.20209437

**Authors:** Paul M. Luethy

## Abstract

The COVID-19 pandemic has been a significant cause of global morbidity and mortality, with evidence suggesting that activities involving heavier breathing, such as singing and exercise, can result in increased risk for disease transmission. The USA Curling Club Nationals is a week-long curling tournament to determine the men’s and women’s club-level champions. The 2020 tournament took place March 7-14 at the Potomac Curling Club in Laurel, MD, and featured teams from across the United States. Preventative measures, such as increased cleaning and disinfection of surfaces, single use and disposable food containers, and canceling traditional event banquets were implemented. Despite these measures, players, coaches, officials, volunteers, and spectators contracted the virus as a result of participation in the event. We surveyed participants to assess total positivity, potential days of transmission, and the burden of symptoms experienced among the participants. We found that 55.6% of all participants reported experiencing symptoms consistent with COVID-19, with nearly all experiencing more than one symptom. Although most participants’ symptoms resolved quickly, 9.6% of all participants experienced symptoms for at least one month and 12.6% of all participants reported taking at least 30 days before they felt they had returned to normal. As a result of this study, we believe curling tournaments have the potential to be high-risk events for the transmission of COVID-19. Further infection prevention measures that were not yet publicly implemented at the time of this tournament may be an effective method of lowering transmission risk, although further research is required.

## Introduction

The Coronavirus Infectious Disease 2019 (COVID-19) pandemic has resulted in the deaths of greater than 200,000 United States citizens and over 1 million deaths worldwide (1). Transmitted through the inhalation of droplets containing the SARS-CoV-2 virus, emerging evidence suggests that the virus may spread through both large droplets that fall to the ground quickly and small droplets that stay suspended in the air longer that may travel greater distances (2). Multiple epidemiological reports have been published by the US Centers for Disease Control and Prevention (CDC) establishing that certain types of activities result in increased spread of COVID-19. Perhaps the most well-known study is that of a 61 person choir where 32 members were confirmed and 20 were likely infected (3), likely due to the indoor nature of the event and the increased number of droplets released due to singing. Gym environments where heavier breathing results in the release of more droplets have also been shown to act as areas of high risk (4). Recently, mask use has been shown to be one important factor to preventing the spread of infection (5) along with other preventative measures.

The USA Curling Club Nationals is a weeklong curling tournament to determine the men’s and women’s club-level champions. The event is organized and sanctioned by the USA Curling Association (USCA). Men’s and women’s teams from each of the 10 regional curling associations compete in a round robin tournament followed by playoffs to decide the champions. The 2020 Club Nationals were held at the Potomac Curling Club in Laurel, Maryland, from March 7-14, with teams from the states of Alaska, Arizona, Colorado, Illinois, New Jersey, Minnesota, Michigan, Washington, North Dakota, and Wisconsin. In an effort to reduce the chance of contracting COVID-19 during the tournament, many changes in practice were made that were thought to be effective at that time in the pandemic. These changes included cleaning of surfaces and rock handles between uses, encouraging use of hand sanitizer, and limiting use of cups and snacks to single-use disposable options. Major event changes included canceling the opening ceremony, offering a limited closing banquet, and discouraging the traditional pre- and post-game handshakes. Although self-limited in gathering size by nature, an official order limiting gatherings larger than 250 people was not put into effect by the state of Maryland until March 12, 2020 (6). On March 18, 2020, the USCA informed participants, volunteers, and spectators that an individual who participated in the tournament had tested positive for COVID-19 (7). On March 27, 2020, the USCA announced that additional participants had tested positive for COVID-19 (8). However, no formal investigation has yet been performed to evaluate the total level of transmission that occurred.

In this study, we surveyed the players, coaches, officials, volunteers, and spectators who attended the event to assess total positivity, potential days of transmission, and the burden of symptoms experienced among the participants. Following an 85% response rate, we found that 55.6% of all participants reported experiencing symptoms consistent with COVID-19, with nearly all experiencing more than one symptom. Although most participants’ symptoms resolved quickly, 9.6% of all participants experienced symptoms for at least one month and 12.6% of all participants reported taking at least 30 days before they felt they had returned to normal. In summary, curling tournaments are at risk for developing into COVID-19 spreader events. Modifications to curling facilities and gameplay (such as increased ventilation, reduced occupancy, and requiring masks), as suggested by current USCA and public health guidelines, are likely required to help prevent transmission in comparison to cleaning protocols alone.

## Methods

This study was submitted to and approved by the University of Maryland, Baltimore (UMB) Institutional Review Board (IRB) as Exempt under protocol number HP-00092884. Study data were collected and managed using REDCap electronic data capture tools hosted at the University of Maryland, Baltimore (9,10). REDCap (Research Electronic Data Capture) is a secure, web-based software platform designed to support data capture for research studies, providing 1) an intuitive interface for validated data capture; 2) audit trails for tracking data manipulation and export procedures; 3) automated export procedures for seamless data downloads to common statistical packages; and 4) procedures for data integration and interoperability with external sources. REDCap software was utilized to build the data collection survey, distribute the survey, securely collect responses, and generate a database. Participants were given the opportunity to submit their responses for a period of two weeks (September 1-13, 2020) before data collection was closed. All data was dissociated from the participant’s unique link such that all responses were anonymous.

Participants were eligible for the survey if they met the following criteria: 1) 18 years of age or older but not older than 88 years of age and 2) were a player, coach, official, volunteer, or spectator. The study size was determined to be 187 individuals, which is the total combined number of participants. The following variables were assessed through the survey: a) participant type (player/alternate/coach; official/ice crew/volunteer; spectator); b) days the participant attended the event; c) gender; d) age group (18-34; 35-54; 55-88 years old); e) whether the participant experienced COVID-19 related symptoms; f) which specific symptoms were experienced; g) was the participant hospitalized; h) length of COVID-19 related symptoms; i) length of time until participant was able to resume normal activities; j) whether the participant received a viral test for COVID-19 and its result; k) whether the participant received an antibody test for COVID-19 and its result; and l) did a close contact become symptomatic or test positive for COVID-19 following the individual’s participation in the tournament. Items f-i were only assessed if the participant answered “yes” to item e.

Statistical methods were descriptive and were completed in R version 4.0.2. Number and percents were described for categorical variables, and mean +/- standard deviation (SD) were described for continuous variables. Violin plots were generated using the ggplot2 package (11) and bar plots and heatmaps were created in the base package of R version 4.0.2 (12). For participants who entered 300 days to indicate they had not yet returned to normal activities, we reassigned that number to 175 days for continuous and graphical representations, as that is the approximate number of days between the event and when the survey was administered.

## Results

159 (85.0%) participants responded to the survey (Table 1). Volunteers/officials who responded to the survey were older (34.7% 55-88 years of age) and more likely to be male (59.7%) compared to players/coaches (24.4% and 50%, respectively).

**Table 1.**
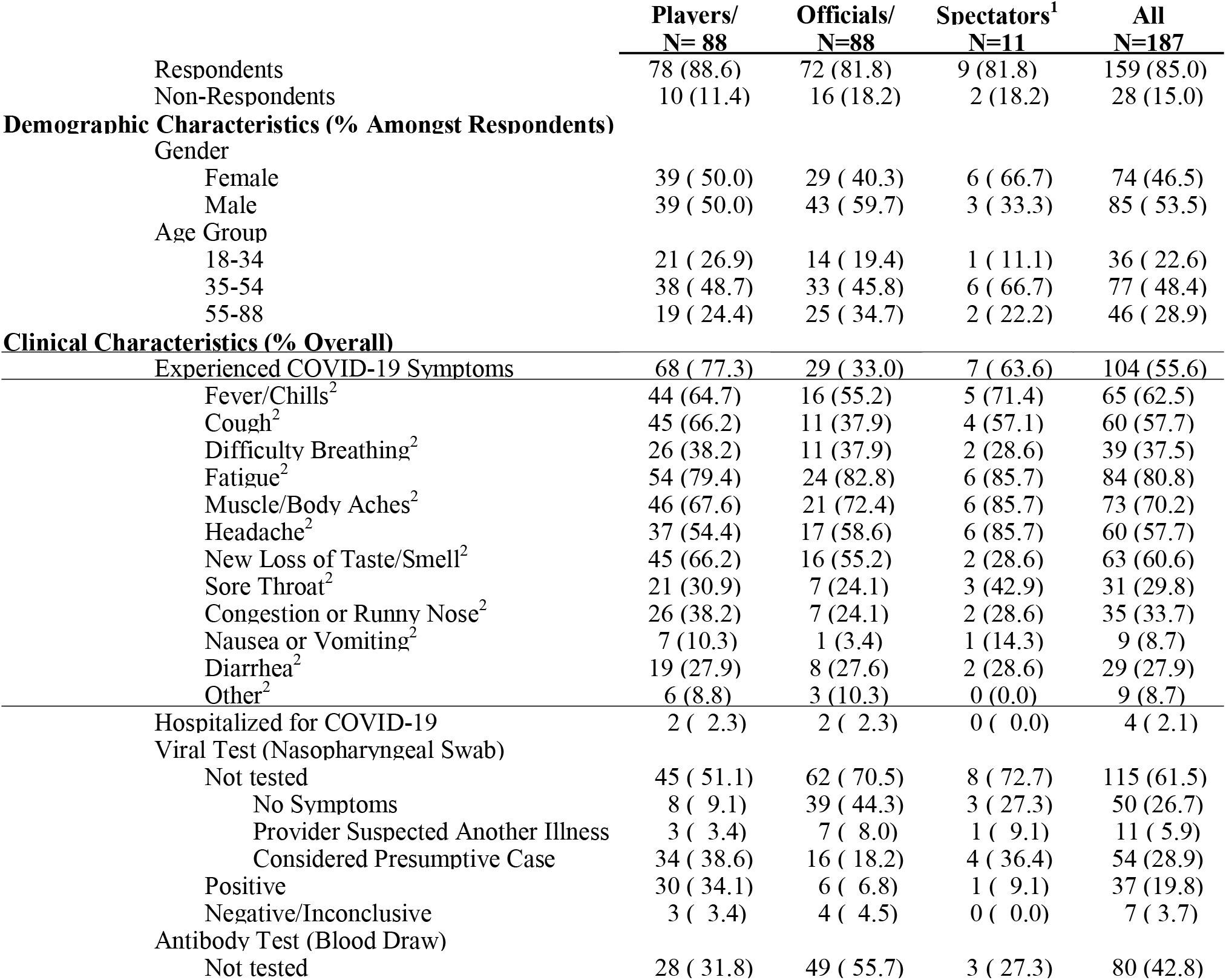
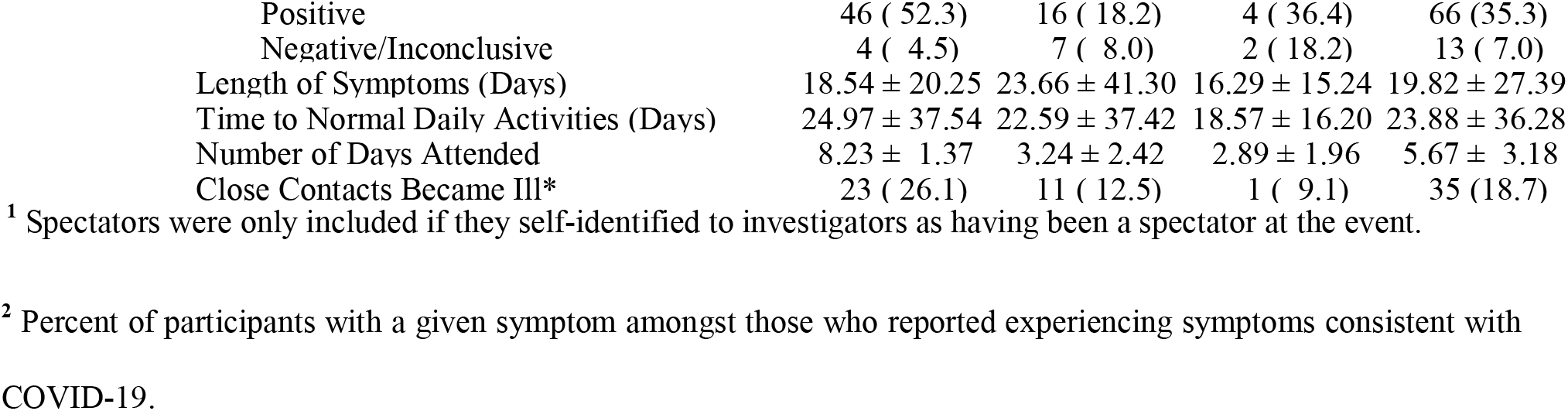
Demographic and clinical characteristics for players/coaches, officials/volunteers, spectators, and all known participants at 2020 US Club Nationals. N(%) for categorical measures and Mean +/- SD for continuous measures. Percents are calculated amongst respondents for demographics and from overall for other categorical variables, except for symptom description, where they are only described amongst participants who reported experiencing COVID-19 symptoms.

|  | Players/<br>N= 88 | Officials/<br>N=88 | Spectators<sup>1</sup><br>N=11 | All<br>N=187 |
| --- | --- | --- | --- | --- |
| Respondents | 78 (88.6) | 72 (81.8) | 9 (81.8) | 159 (85.0) |
| Non-Respondents | 10 (11.4) | 16 (18.2) | 2 (18.2) | 28 (15.0) |
| **Demographic Characteristics (% Amongst Respondents)** |  |  |  |  |
| Gender |  |  |  |  |
| Female | 39 ( 50.0) | 29 ( 40.3) | 6 ( 66.7) | 74 (46.5) |
| Male | 39 ( 50.0) | 43 ( 59.7) | 3 ( 33.3) | 85 ( 53.5) |
| Age Group |  |  |  |  |
| 18-34 | 21 ( 26.9) | 14 ( 19.4) | 1 ( 11.1) | 36 ( 22.6) |
| 35-54 | 38 ( 48.7) | 33 ( 45.8) | 6 ( 66.7) | 77 ( 48.4) |
| 55-88 | 19 ( 24.4) | 25 ( 34.7) | 2 ( 22.2) | 46 ( 28.9) |
| **Clinical Characteristics (% Overall)** |  |  |  |  |
| Experienced COVID-19 Symptoms | 68 ( 77.3) | 29 ( 33.0) | 7 ( 63.6) | 104 (55.6) |
| Fever/Chills<sup>2</sup> | 44 (64.7) | 16 (55.2) | 5 (71.4) | 65 (62.5) |
| Cough<sup>2</sup> | 45 (66.2) | 11 (37.9) | 4 (57.1) | 60 (57.7) |
| Difficulty Breathing<sup>2</sup> | 26 (38.2) | 11 (37.9) | 2 (28.6) | 39 (37.5) |
| Fatigue<sup>2</sup> | 54 (79.4) | 24 (82.8) | 6 (85.7) | 84 (80.8) |
| Muscle/Body Aches<sup>2</sup> | 46 (67.6) | 21 (72.4) | 6 (85.7) | 73 (70.2) |
| Headache<sup>2</sup> | 37 (54.4) | 17 (58.6) | 6 (85.7) | 60 (57.7) |
| New Loss of Taste/Smell<sup>2</sup> | 45 (66.2) | 16 (55.2) | 2 (28.6) | 63 (60.6) |
| Sore Throat<sup>2</sup> | 21 (30.9) | 7 (24.1) | 3 (42.9) | 31 (29.8) |
| Congestion or Runny Nose<sup>2</sup> | 26 (38.2) | 7 (24.1) | 2 (28.6) | 35 (33.7) |
| Nausea or Vomiting<sup>2</sup> | 7 (10.3) | 1 (3.4) | 1 (14.3) | 9 (8.7) |
| Diarrhea<sup>2</sup> | 19 (27.9) | 8 (27.6) | 2 (28.6) | 29 (27.9) |
| Other<sup>2</sup> | 6 (8.8) | 3 (10.3) | 0 (0.0) | 9 (8.7) |
| Hospitalized for COVID-19 | 2 ( 2.3) | 2 ( 2.3) | 0 ( 0.0) | 4 ( 2.1) |
| Viral Test (Nasopharyngeal Swab) |  |  |  |  |
| Not tested | 45 ( 51.1) | 62 ( 70.5) | 8 ( 72.7) | 115 (61.5) |
| No Symptoms | 8 ( 9.1) | 39 ( 44.3) | 3 ( 27.3) | 50 (26.7) |
| Provider Suspected Another Illness | 3 ( 3.4) | 7 ( 8.0) | 1 ( 9.1) | 11 ( 5.9) |
| Considered Presumptive Case | 34 ( 38.6) | 16 ( 18.2) | 4 ( 36.4) | 54 (28.9) |
| Positive | 30 ( 34.1) | 6 ( 6.8) | 1 ( 9.1) | 37 (19.8) |
| Negative/Inconclusive | 3 ( 3.4) | 4 ( 4.5) | 0 ( 0.0) | 7 ( 3.7) |
| Antibody Test (Blood Draw) |  |  |  |  |
| Not tested | 28 ( 31.8) | 49 ( 55.7) | 3 ( 27.3) | 80 (42.8) |
| Positive | 46 ( 52.3) | 16 ( 18.2) | 4 ( 36.4) | 66 (35.3) |
| Negative/Inconclusive | 4 ( 4.5) | 7 ( 8.0) | 2 ( 18.2) | 13 ( 7.0) |
| Length of Symptoms (Days) | 18.54 ± 20.25 | 23.66 ± 41.30 | 16.29 ± 15.24 | 19.82 ± 27.39 |
| Time to Normal Daily Activities (Days) | 24.97 ± 37.54 | 22.59 ± 37.42 | 18.57 ± 16.20 | 23.88 ± 36.28 |
| Number of Days Attended | 8.23 ± 1.37 | 3.24 ± 2.42 | 2.89 ± 1.96 | 5.67 ± 3.18 |
| Close Contacts Became Ill* | 23 ( 26.1) | 11 ( 12.5) | 1 ( 9.1) | 35 (18.7) |

<sup>1</sup> Spectators were only included if they self-identified to investigators as having been a spectator at the event.
<sup>2</sup> Percent of participants with a given symptom amongst those who reported experiencing symptoms consistent with COVID-19.

104 (104/187 = 55.6%) of the participants who were known to have attended the event reported experiencing symptoms consistent with COVID-19 (Table 1). There was a marked disparity in percentage of players/coaches who reported having symptoms compared to volunteers/officials who reported having symptoms: 68 (68/88 = 77.3%) of players/coaches who attended the event reported experiencing COVID-19 symptoms compared to 29 (29/88 = 33.0%) of volunteers/officials. The most frequent symptoms reported by participants who experienced COVID-19 symptoms were fatigue (n=84, 80.8%), muscle/body aches (n=73, 70.2%), fever/chills (n=65, 62.5%), new loss of taste or smell (n=63, 60.6%), cough (n=60, 57.7%), headache (n=60, 57.7%), shortness of breath or difficulty breathing (n=39, 37.5%), and congestion or runny nose (n=35, 33.7%). Some participants reported other symptoms (n=9, 8.7%) in an open-ended, optional question including rash/skin conditions (n=4, 3.8%), heart abnormalities (n=2, 1.9%), loss of appetite (n=2, 1.9%), paranoia (n=1, 1.0%), hoarse voice (n=1, 1.0%), lower back pain (n=1, 1.0%), and COVID fingers/toes (n=1, 1.0%). Most participants who were symptomatic reported multiple symptoms (Figure 1). Amongst participants who reported experiencing COVID-19 symptoms, 97.1%, 86.5%, 75.0%, 59.6%, and 45.2% experienced greater than or equal to 2, 3, 4, 5, or 6 symptoms, respectively.

**Figure 1.**
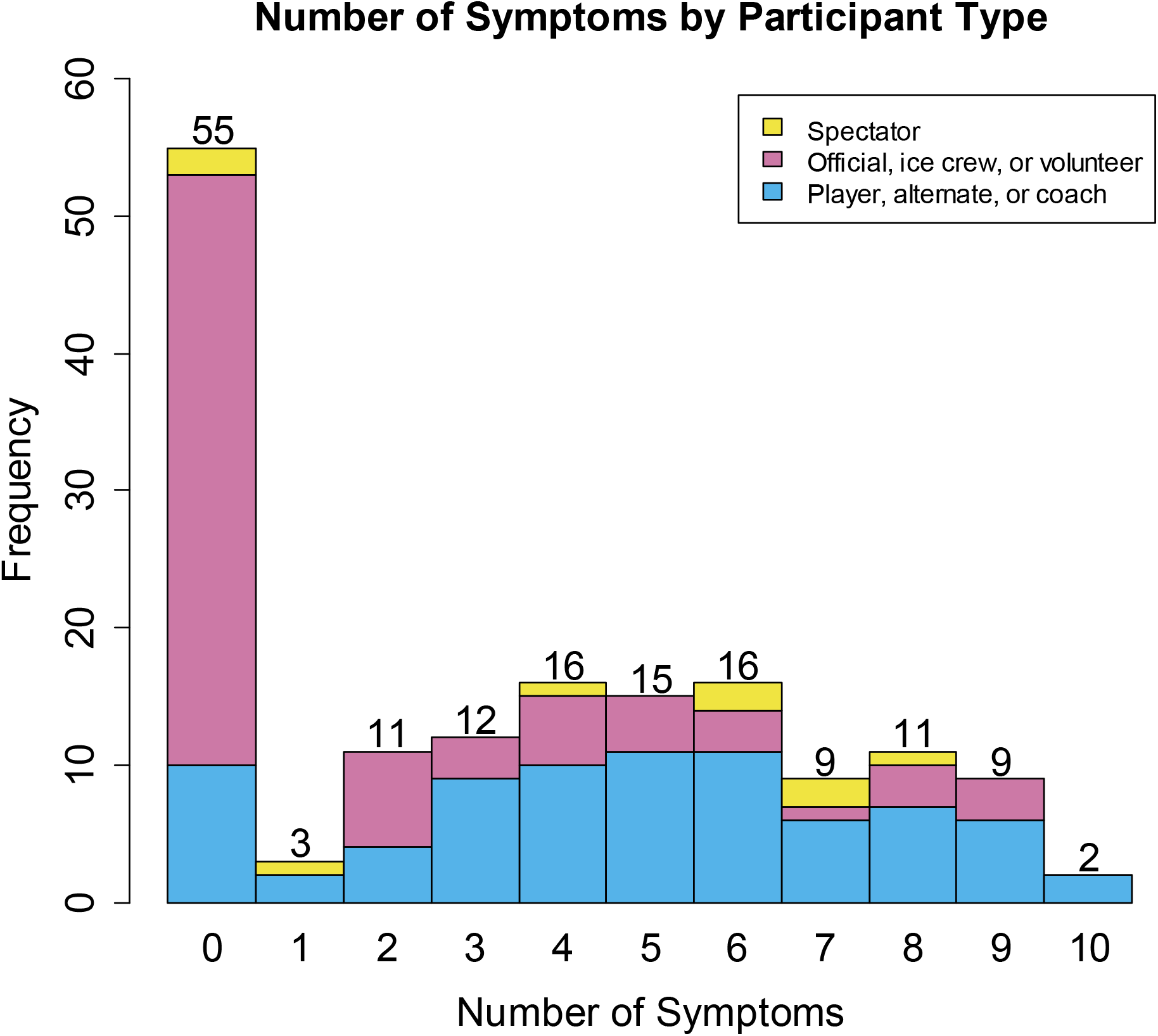
Number of symptoms by participant type. The number of symptoms experienced by each participant type is shown. The total frequency representing the combined numbers from each participant type is displayed above each frequency bar.

Overall, 4 out of 187 (2.1%) participants known to have attended the event were hospitalized (Table 1). 18 (9.6%), 4 (2.1%), 3 (1.6%), and 3 (1.6%) participants who were known to attend the event experienced symptoms for greater than or equal to 30, 60, 90, and 150 days, respectively (Figure 2A). In addition, 23 (12.3%), 13 (7.0%), 6 (3.2%), and 4 (2.1%) participants who were known to attend the event were able to return to normal activities at or later than 30, 60, 90, and 150 days, respectively (Figure 2B). 35 (18.7%) participants reported being concerned that a close contact who did not attend the event became ill after the event (Table 1).

**Figure 2.**
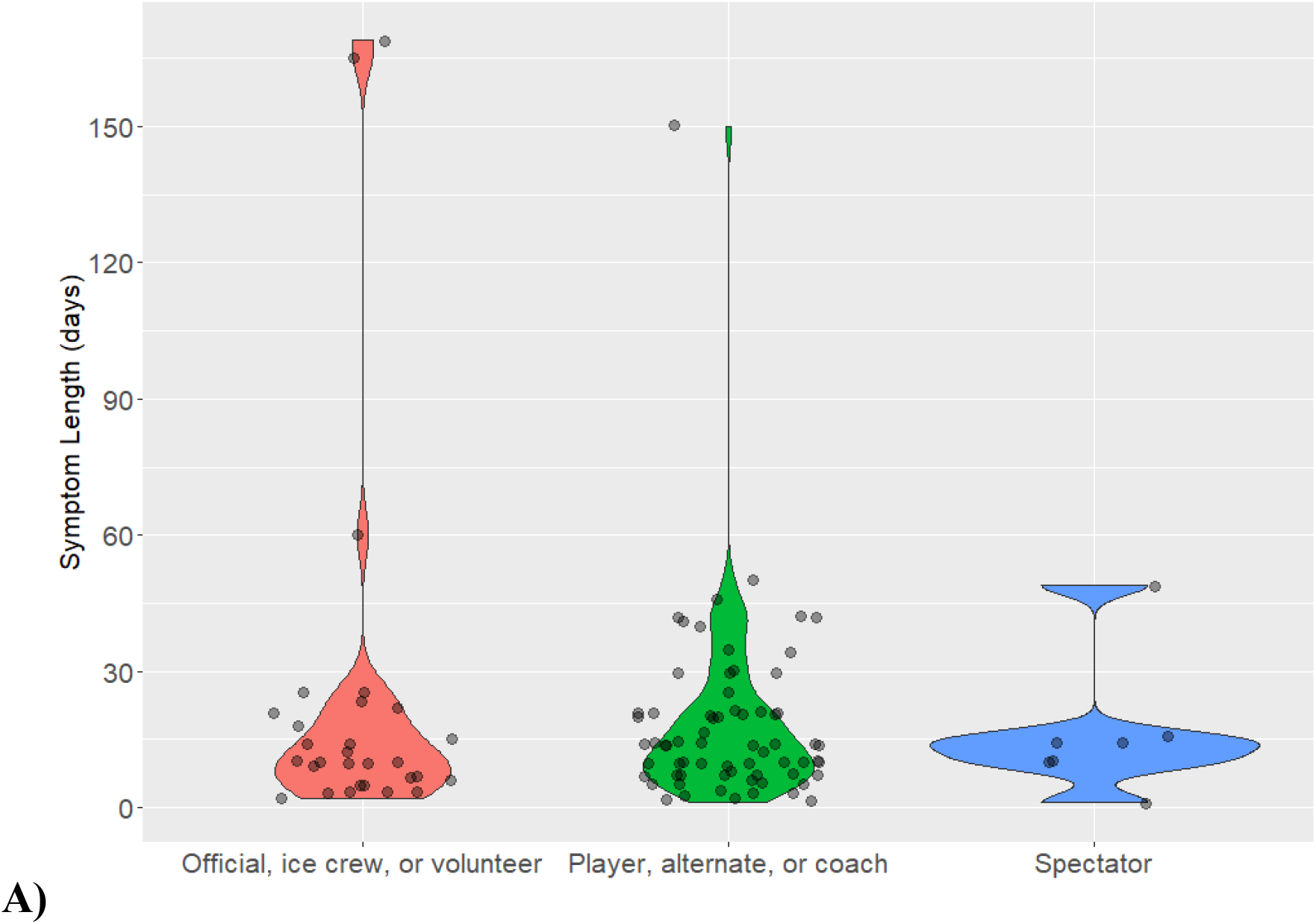
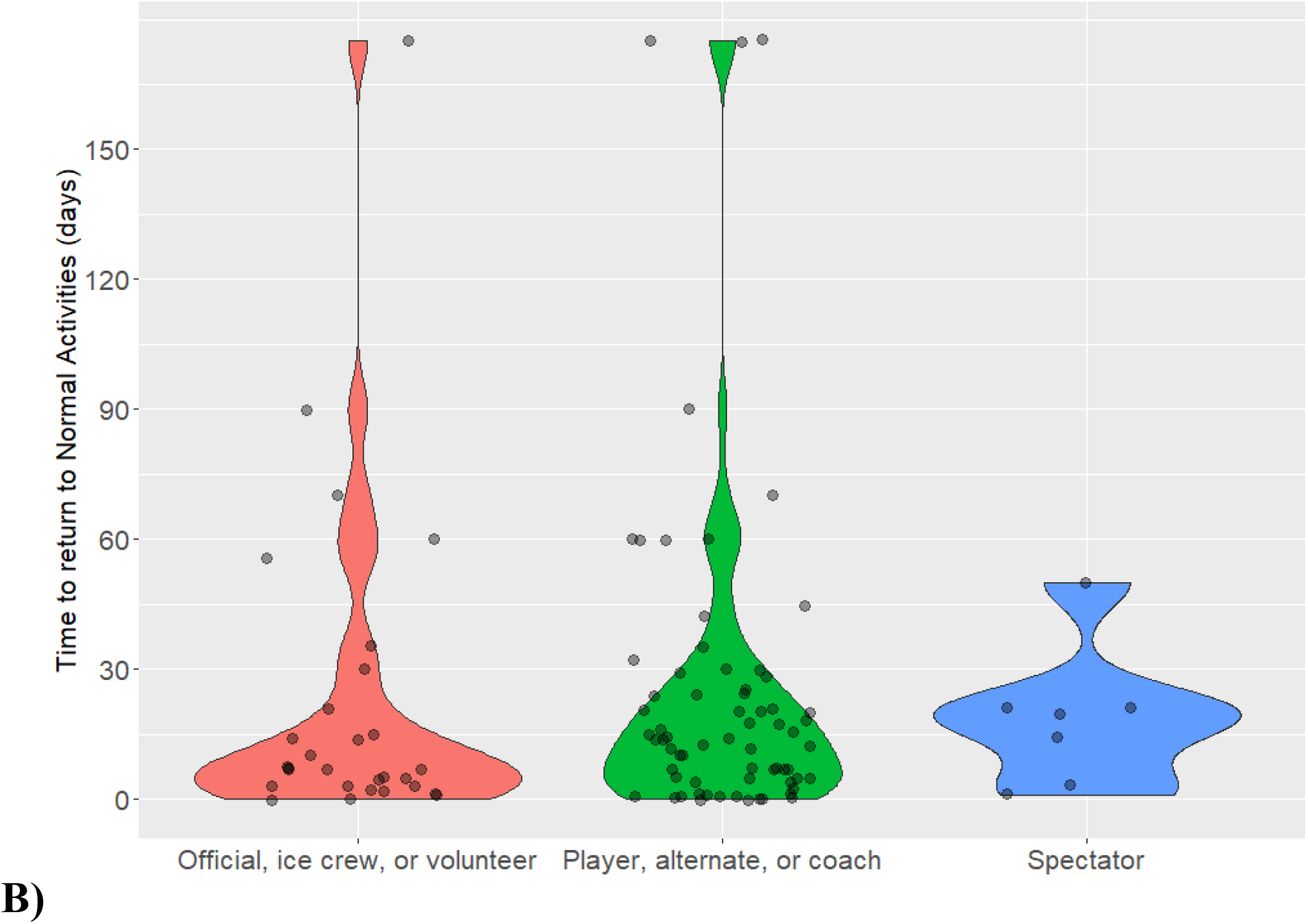
Violin plots depicting **A)** symptom length (days) and **B)** time to return to normal activities (days) by participant type.

Testing was generally not available for all participants in mid-March after the event concluded. Overall, 115 participants (61.5%) were not viral tested by a nasopharyngeal swab, the only type of viral test that was available at the time (Table 1). 50 (26.7%) participants were not viral tested because they did not experience symptoms, 11 (5.9%) were not viral tested because their healthcare provider suspected another illness, and 54 (28.9%) were not tested but were considered a presumptive case by their healthcare provider. 37 (19.8%) of all participants known to have attended the event had positive viral test results while 7 (3.7%) of all participants had negative viral test results (Table 1). Many more participants were able to be tested for antibodies in the months following the event. 66 (35.3%) of all participants known to attend the event tested positive for COVID-19 antibodies while 7 (3.7%) of all participants tested negative or inconclusive for COVID-19 antibodies (Table 1). Table 2 describes the number of respondents by symptoms, viral test result, and antibody test result. For example, amongst participants who reported having COVID-19 symptoms, 22 were not tested by either testing method, 7 were not viral tested but had negative/inconclusive antibody test results, and 36 were viral positive (Table 2). Amongst respondents, 76 (76/159 = 40.6%) participants were positive by viral, antibody, or both tests; 2 (1.1%) participants were negative by both; 14 (7.5%) participants were not viral tested but were negative by an antibody test; and 67 (35.8%) participants were not tested by either method, 45 (67.2%) of whom did not experience any symptoms.

**Table 2.** Number of participants by symptoms, viral test result, and antibody test result.

| **Viral Test Result** | **Antibody Test Result** | **No Symptoms** | **Symptoms** |
| --- | --- | --- | --- |
| Not Tested | Not Tested | 45 | 22 |
|  | Negative/Inconclusive | 4 | 7 |
|  | Positive | 1 | 36 |
| Negative/Inconclusive | Not Tested | 3 | 0 |
|  | Negative/Inconclusive | 1 | 1 |
|  | Positive | 0 | 2 |
| Positive | Not Tested | 0 | 10 |
|  | Negative/Inconclusive | 0 | 0 |
|  | Positive | 1 | 26 |

Figure 3 depicts heat maps describing the days that symptomatic (Figure 3A) and asymptomatic (Figure 3B) volunteers attended. Each row designates a specific volunteer, where a blue rectangle above a date denotes that the volunteer attended that day. For example, symptomatic volunteer 1 (bottom of Figure 3A) attended the event every day from Sunday, March 8 through Sunday, March 15. Symptomatic volunteers tended to attend later in the week and for more days (Figure 3A), while asymptomatic volunteers tended to attend earlier in the week and for fewer days (Figure 3B). In particular, symptomatic volunteers attended an average of 4.4 days and 86.2% attended Thursday or later, while asymptomatic volunteers attended an average of 2.5 days and only 32.6% attended Thursday or later. Plots were also created for the players/coaches (data not shown). However, because these individuals all attended nearly every day, no pattern relating to development of symptoms and days attended was evident.

**Figure 3.**
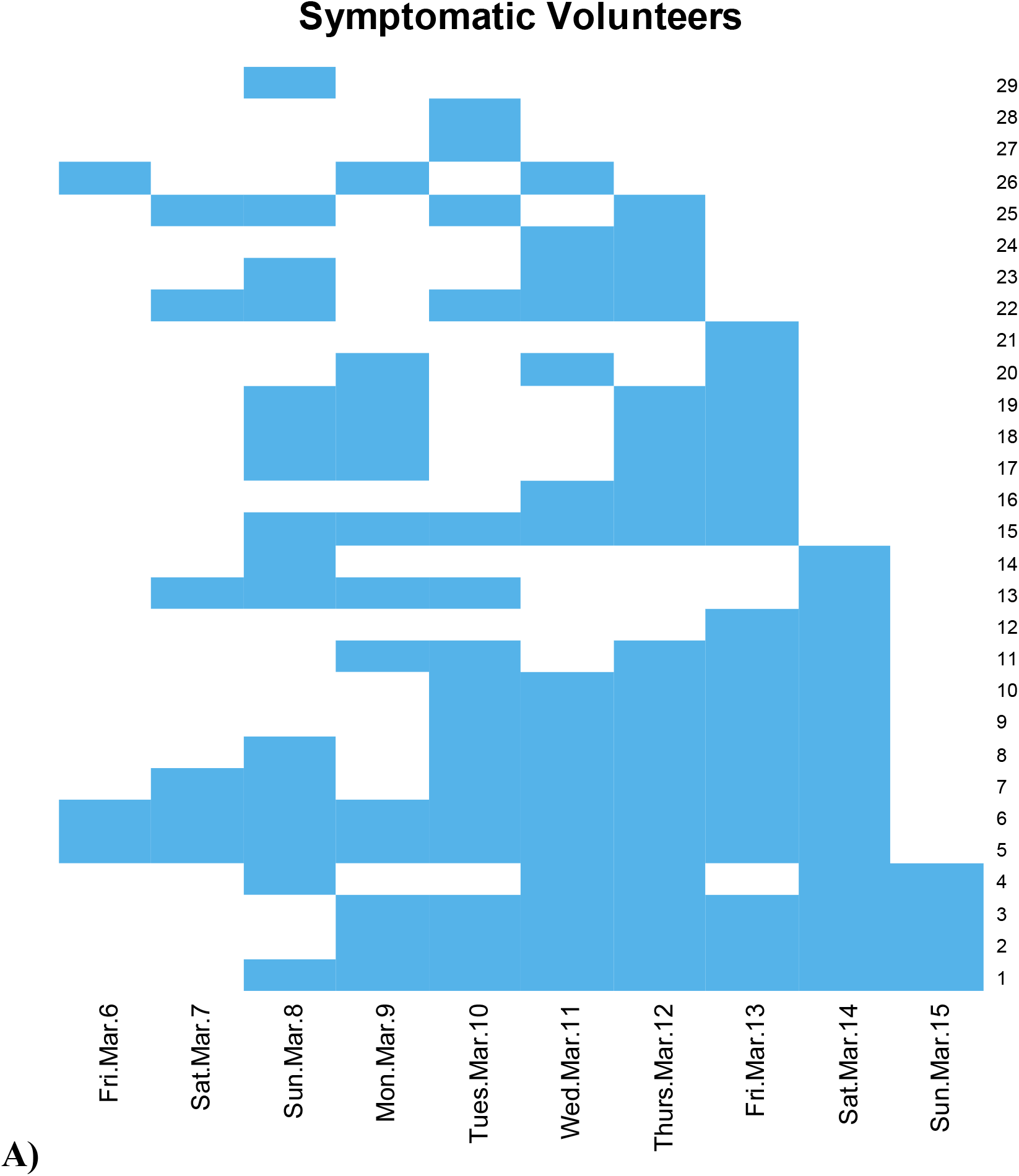
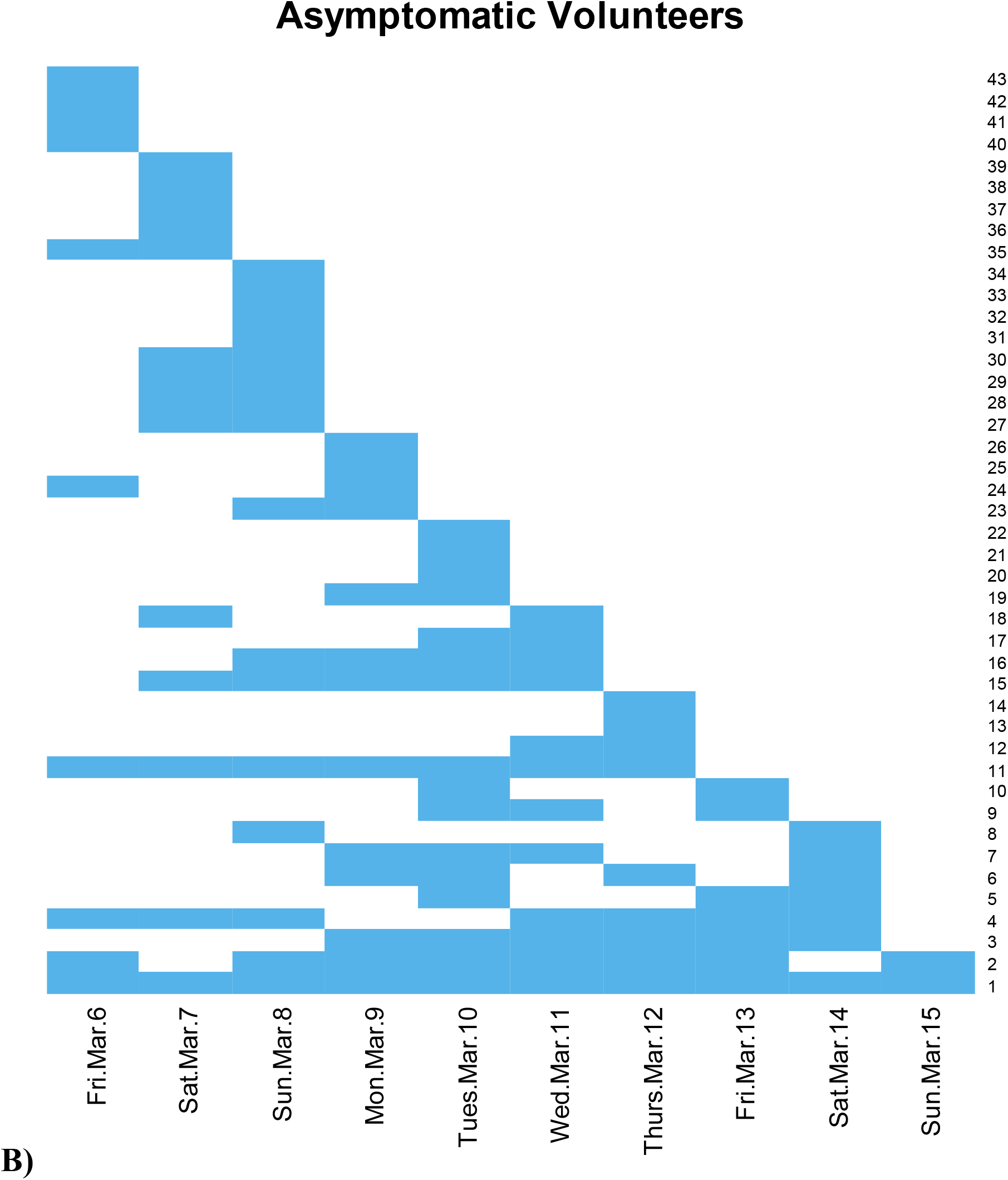
Heat maps depicting the days on which volunteers/officials attended the event by volunteers/officials who **A)** reported experiencing COVID-19 symptoms and **B)** did not report experiencing COVID-19 symptoms. Blue squares represent days the volunteer/official was present and the y-axis represents a unique individual.

## Discussion

According to the CDC, gatherings are a significant risk for COVID-19 transmission (13). The 2020 USA Curling Club Nationals tournament involved a total of 187 known players, coaches, officials, volunteers, and spectators across a nine-day period. During the tournament, not all of these individuals were present at once, with approximately 40 players and officials on the ice at once and approximately 40 players, coaches and volunteers in the warm room during a normal game session. These repeated gatherings over the course of the tournament proved to be sufficient for transmission, as we observed that a large number of participants (n= 104, 55.6%) at the event experienced symptoms consistent with COVID-19. Not surprisingly, there was a large disparity between the percentage of players/coaches (n= 68, 77.3%) who experienced symptoms and volunteers/officials (n=29, 33.0%) who experienced symptoms. It is likely that the length of time at the event played a big factor here as most players/coaches attended every day of practice/competition (8.23 days attended, on average) while volunteers/officials spent less time at the event (3.24 days attended, on average). Indeed, even amongst volunteers, volunteers reporting symptoms tended to attend more frequently and later in the week than volunteers reporting no symptoms (Figure 3).

In order to estimate when the virus may have been widely circulating, we assessed symptomatic volunteers whose last day was early in the week (Figure 3A). In particular, we looked at four symptomatic volunteers (26 through 29). Symptomatic volunteer 29 only attended on Sunday, March 8, reported having fever/chills and a headache and was not tested using either method because their healthcare provider suspected another illness. Symptomatic volunteers 27 & 28 only attended on Tuesday, March 10. One had cough, shortness breath/difficulty breathing, fatigue, muscle/body aches, and was not tested via either method but was considered a presumptive case by their healthcare provider. The other volunteer had muscle/body aches, headache, sore throat, and was tested for both with a negative/inconclusive result for both. Symptomatic volunteer 26 attended three days, with their last day on Wednesday, March 11. This individual reported fever/chills, body aches, loss of taste/smell, cough, shortness breath/difficulty breathing, fatigue, headache, sore throat, and was not tested via either method but was considered a presumptive case by their healthcare provider. In comparison, symptomatic volunteers 22 through 25 last attended on Thursday, March 12. All four had multiple symptoms consistent with COVID-19. One tested positive for both viral and antibody tests, while the other three were not viral tested but were considered presumptive cases, two of whom later tested positive for antibodies. It therefore appears that COVID-19 was widely circulating by Thursday, March 12, but may have been circulating less widely earlier than that date.

This survey had a high rate of response; 159 out of 187 participants (85%) known to have attended the event responded to the survey. It is noted that there is likely to be selection bias in respondents in that participants who had symptoms consistent with COVID-19 were more likely to respond to the survey than participants who did not have symptoms. However, to address this possibility, we reported percentages of participants with COVID-19 symptoms, hospitalization rates, and test rates using the denominator of known participants (187) instead of the number of participants who responded (159) to be more conservative in our estimates. Our estimates therefore reflect a “best case” scenario in that we assume that non-respondents did not have positive test results or symptoms consistent with COVID-19. It is possible that several of the remaining 28 participants that did not respond to the survey experienced symptoms of COVID-19 and/or tested positive by a viral or antibody test. However, we do not believe that these additional responses would dramatically alter the results we observed.

A particular limitation of this study was the lack of testing—particularly viral testing, which is considered to be the gold standard for diagnosing COVID-19—available after the conclusion of the event. Due to this limitation, we have mainly focused our descriptions on the burden of symptoms in participants. In addition, many participants (n=35, 18.7%) reported being concerned that one or more close contacts who did not attend the event became symptomatic or tested positive after the event. Although this question was meant to assess potential secondary cases, another limitation of this study is the authors’ lack of resources and expertise to complete contact tracing after the event. Indeed, it would be a large task as many participants flew home after being exposed at the event. Finally, this study did not assess whether symptomatic volunteers were stationed in the ice shed or the warm room in relation to their volunteer assignment. This question could have helped to determine which environment represents a larger transmission risk. However, this likely would have been difficult to assess, as some volunteers often spent time in the ice shed and warm room as a part of their duties. We believe that both environments were potential transmission areas as evidenced by the 77.3% of players and coaches who developed COVID-19 symptoms and spent significant time on the ice and in the warm room.

In conclusion, we observed that curling tournaments can pose a significant risk for transmission of COVID-19, especially to players and volunteers who have repeated exposure to infected individuals. Importantly, the spread of infection was observed even with stringent cleaning protocols and modifications to gameplay and tournament traditions in place. The USCA has since released guidelines detailing modifications to facility operations (such as closing locker rooms), gameplay (using a single sweeper), limiting or removing post-game socializing, and requiring masks both on and off the ice (14). Further research is required to determine whether these guidelines are sufficient to prevent spread of COVID-19 within a curling facility.

## Supporting information

Conflict of Interest Disclosure

STROBE checklist

## Data Availability

Datasets are available upon request to the corresponding author and following approval from the University of Maryland Baltimore IRB.

## Acknowledgements

We would like to thank Linda Kirkman and Kerry Hadiaris for their help in designing survey questions and advertising to potential participants. Lastly, we would like to thank the USCA and Potomac Curling Club for their help in recruiting participants for this study. This study was not supported by any external funding sources.

## References

1. Mortality Analyses [Internet]. Johns Hopkins Coronavirus Resource Center. [cited 2020 Sep 28]. Available from: https://coronavirus.jhu.edu/data/mortality

2. Jayaweera M, Perera H, Gunawardana B, Manatunge J. Transmission of COVID-19 virus by droplets and aerosols: A critical review on the unresolved dichotomy. Environ Res. 2020 Sep;188:109819.

3. Hamner L. High SARS-CoV-2 Attack Rate Following Exposure at a Choir Practice — Skagit County, Washington, March 2020. MMWR Morb Mortal Wkly Rep [Internet]. 2020 [cited 2020 Sep 28];69. Available from: https://www.cdc.gov/mmwr/volumes/69/wr/mm6919e6.htm

4. Jang S, Han SH, Rhee J-Y. Cluster of Coronavirus Disease Associated with Fitness Dance Classes, South Korea - Volume 26, Number 8—August 2020 - Emerging Infectious Diseases journal - CDC. [cited 2020 Sep 28]; Available from: https://wwwnc.cdc.gov/eid/article/26/8/20-0633_article

5. Hendrix MJ. Absence of Apparent Transmission of SARS-CoV-2 from Two Stylists After Exposure at a Hair Salon with a Universal Face Covering Policy — Springfield, Missouri, May 2020. MMWR Morb Mortal Wkly Rep [Internet]. 2020 [cited 2020 Sep 28];69. Available from: https://www.cdc.gov/mmwr/volumes/69/wr/mm6928e2.htm

6. OFFICE OF GOVERNOR LARRY HOGAN. Governor Larry Hogan - Official Website for the Governor of Maryland: COVID-19 Pandemic: Orders and Guidance [Internet]. [cited 2020 Sep 28]. Available from: https://governor.maryland.gov/covid-19-pandemic-orders-and-guidance/

7. Potomac Curling Club on Twitter [Internet]. Twitter. [cited 2020 Sep 28]. Available from: https://twitter.com/curldc/status/1240278753365970944

8. USA Curling Provides Update On Club Nationals [Internet]. Team USA. [cited 2020 Sep 28]. Available from: https://www.teamusa.org:443/USA-Curling/Features/2020/March/27/USA-Curling-Provides-Update-On-Club-Nationals

9. Harris PA, Taylor R, Thielke R, Payne J, Gonzalez N, Conde JG. Research electronic data capture (REDCap)—A metadata-driven methodology and workflow process for providing translational research informatics support. Journal of Biomedical Informatics. 2009 Apr 1;42(2):377–81.

10. Harris PA, Taylor R, Minor BL, Elliott V, Fernandez M, O’Neal L, et al. The REDCap consortium: Building an international community of software platform partners. Journal of Biomedical Informatics. 2019 Jul 1;95:103208.

11. Wickham, Hadley. ggplot2: Elegant Graphics for Data Analysis [Internet]. Springer-Verlag New York; 2016 [cited 2020 Sep 28]. Available from: https://ggplot2.tidyverse.org/

12. R Core Team. R: A language and environment for statistical computing. [Internet]. Vienna, Austria: R Foundation for Stastistical Computing; 2020 [cited 2020 Sep 28]. Available from: https://www.r-project.org/

13. CDC. Communities, Schools, Workplaces,& Events [Internet]. Centers for Disease Control and Prevention. 2020 [cited 2020 Sep 28]. Available from: https://www.cdc.gov/coronavirus/2019-ncov/community/large-events/considerations-for-events-gatherings.html

14. COVID-19 Updates [Internet]. Team USA. [cited 2020 Sep 28]. Available from: https://www.teamusa.org:443/USA-Curling/COVID-19-Updates

